# Diffusion-limited O_2_ release in human kidneys perfused with stored blood

**DOI:** 10.1101/2023.05.07.23289584

**Authors:** Richard Dumbill, Julija Rabcuka, Simon Knight, James Hunter, John Fallon, Daniel Voyce, Jacob Barrett, Matt Ellen, Annemarie Weissenbacher, Tetuko Kurniawan, Slawomir Blonski, Piotr Korczyk, Rutger Ploeg, Constantin Coussios, Peter Friend, Pawel Swietach

## Abstract

A central dogma in physiology is that oxygen release at tissues is not diffusion-limited because gas exchange at capillaries is rapid. This assertion has influenced clinical care, which focuses on optimising oxygen delivery through improving blood flow and oxygen content, rather than oxygen unloading from red blood cells (RBCs). Since storage of blood causes profound changes that slow oxygen release from RBCs, transfusions could compromise tissue respiration. We investigated this in transplant human kidneys normothermically perfused with stored blood. During perfusions, renal respiration was measured from blood gases and RBCs were analysed for oxygen-unloading kinetics. Respiratory rate did not correlate significantly with the standard definition of oxygen delivery based on blood flow and oxygen content. However, a strong correlation was obtained after introducing a factor describing oxygen release from RBCs. Oxygen release to tissues can become diffusion-limited with transfused blood, and therefore the kinetic quality of RBCs should be considered.

## INTRODUCTION

Oxygen delivery to respiring cells is a fundamentally important physiological process and a primary concern in intensive care and transfusion medicine. The canonical view^1^ holds that gas exchange in capillaries is rapid, such that oxygen partial pressures (P_O2_) equilibrate between blood and tissue by the time oxygen-carrying red blood cells (RBCs) reach the venous end of capillaries. For resting cardiac output, equilibrium between alveolar and capillary P_O2_ is achieved around 1/3 down the length of pulmonary capillaries, which gives considerable reserve with higher flows^2,3^. Even with strenuous exercise, equilibration is expected, except in some elite athletes who may show a phenomenon of arterial hypoxaemia because of incomplete oxygen loading^4^. Similarly, rapid equilibration is assumed in systemic capillaries, where oxygen is offloaded. Consequently, even moderate changes to the rate of oxygen unloading from RBCs are predicted to have minimal impact on the total amount of oxygen released, because equilibrium is expected eventually. Under most circumstances, oxygen delivery to tissues is, therefore, considered a perfusion-limited process that can be modelled as the product of blood flow (Q), blood haemoglobin concentration ([Hb]) and arterial HbO_2_ saturation (S_A_), but independent of the kinetics of oxygen unloading from RBCs. A perfusion-limited system is intuitively desirable for a process as important as oxygen delivery because it ensures complete exchange, which means that the only limiting factor for oxygen supply to tissues is the rate at which oxygen is delivered by arterial blood. The respiratory rate, set by the metabolic demands of tissue, will extract the required amount of oxygen from blood during its capillary transit, provided that the transit time of RBCs along capillaries is sufficiently long^5^. Oxygen extraction reduces venous HbO_2_ saturation (S_v_) proportionately, but its effect on venous P_O2_ will depend on the shape of the Hb-O_2_ dissociation curve. For example, depletion of 2,3-diphosphoglycerate (2,3-DPG) in RBC cytoplasm will shift this curve to the left which means that for a given O_2_ extraction, venous P_O2_ should fall further.

The assumption that O_2_ exchange in systemic capillaries is not diffusion-limited has had major implications on clinical care. For example, interventions recommended for raising oxygen delivery^6^ include increasing blood flow (e.g. by positive inotropes), raising haemoglobin (e.g. transfusion) or ensuring full oxygen saturation of arterial blood (e.g. ventilation on oxygen), but not stimulating gas-handling kinetics by RBCs. The presumed rapidity of O_2_ unloading from blood implies that disturbances to RBCs are less likely to affect gas exchange because of spare capacity. An example of such a cellular disturbance takes place in blood under storage^7,8^. Stored RBCs undergo biochemical and morphological changes that collectively cause oxygen unloading to become slower^9^. If, however, this functional attrition was within the capillary transit time, then overall oxygen delivery, and hence respiratory rate, should not be affected. Conventionally, it is therefore assumed that the storage lesion does not compromise the efficacy of blood transfusions^10^. This assertion is consistent with human and animal studies showing that the extent of storage lesion, quantified in terms of storage duration, has no negative impact on transfusion outcomes^11-13^, although these inferences have been critiqued on the basis that storage time is not an accurate proxy of actual blood quality^14^.

Perfusion-limited oxygen delivery underpins Early Goal-Directed Therapy (EGDT)^15-18^, which refers to the protocolised management of patients with sepsis. Various haemodynamic variables, including central venous oxygen saturation, are used to guide administration of fluids, packed red cells, inotropes, and vasopressors to increase blood flow, increase haemoglobin concentration, and reduce the oxygen extraction ratio. Whilst early studies were promising^15^, a meta-analysis of three pivotal randomised controlled trials has suggested that there is little benefit associated with EGDT and its practice is not widespread^19,20^. Indeed, systematic review and meta-analysis of studies specifically considering transfusion threshold in this patient population has shown no benefit from transfusion^21^, and the current International Guidelines for Management of Sepsis and Septic Shock recommends a restrictive transfusion policy^22^. A potential reason for the failure of EGDT (and particularly of its transfusion component) to show more clinical benefit could be that under extreme conditions, oxygen release from RBCs may become diffusion-limited. Failure of an increase in haemoglobin concentration to improve outcome when oxygen extraction is high could be explained by diffusion-limited release of oxygen from the transfused cells.

Recently, our group has developed single-cell oxygen saturation imaging to demonstrate that oxygen unloading from RBCs is considerably slower than previous estimates^23^, based largely on the kinetics of oxygen release from acellular haemoglobin solutions^24-28^. The additional delay imposed by RBCs is due to slow diffusivity across a dense cytoplasm, causing diffusional delays even over the short distance of RBC thickness. Consequently, changes in cell shape that expand diffusion pathlength may have profound impact on gas exchange kinetics, as shown in diseases such as hereditary spherocytosis^23^ and in stored blood^9,29^. Critically, the progression of kinetic dysfunction can be slowed under hypoxic storage^29^ and reversed by biochemical rejuvenation^9^. However, the impact of slower-than-expected O_2_ unloading from RBCs on tissue respiration in perfused organs is unclear, because it may still be possible for blood oxygen to equilibrate during capillary transit lasting a few seconds^30,31^ even at the measured unloading rate in RBCs *ex vivo*. To address this question, we analysed RBCs used during prolonged-duration Normothermic Machine Perfusion of the kidney (NMP-K)^32,33^, conducted as part of a phase 1 trial at the Churchill Hospital, Oxford, to evaluate the safety and efficacy of NMP-K prior to deceased-donor renal transplantation (Normothermic Machine Perfusion Phase 1, NKP1).

There are several reasons why the kidney is a suitable organ for testing whether gas-handling by RBCs can result in diffusion-limited oxygen uptake by tissues. Firstly, renal blood flow and respiratory rate are proportional under physiological conditions, thus any discrepancy from this relationship could indicate diffusion-limitation (**Figure 1A**). This physiological coupling arises because the blood flowing to the kidney carries both the substrate for tubular reabsorption and the oxygen needed for respiration to power these active transport processes. Consequently, an increase in blood flow is expected to raise oxygen delivery and demand in tandem. Secondly, the kidney’s physiological function can be monitored to control for tissue viability using indices such as urinary sodium release or creatine clearance. Thirdly, renal oxygen extraction under physiological conditions is modest, at 10-15%, which means that the supply of oxygen from blood is not normally limiting and venous partial pressures rarely fall to low levels. This also means that P_O2_ gradients are relatively small in the kidney, thus any diffusion-limited process is more likely to emerge. Compared to many other organs, there is less variation between individuals in terms of kidney vascular function, which can also be controlled pharmacologically and monitored during perfusions. Finally, kidney transplants are relatively common and therefore a readily available organ for studies.

**Figure 1:**
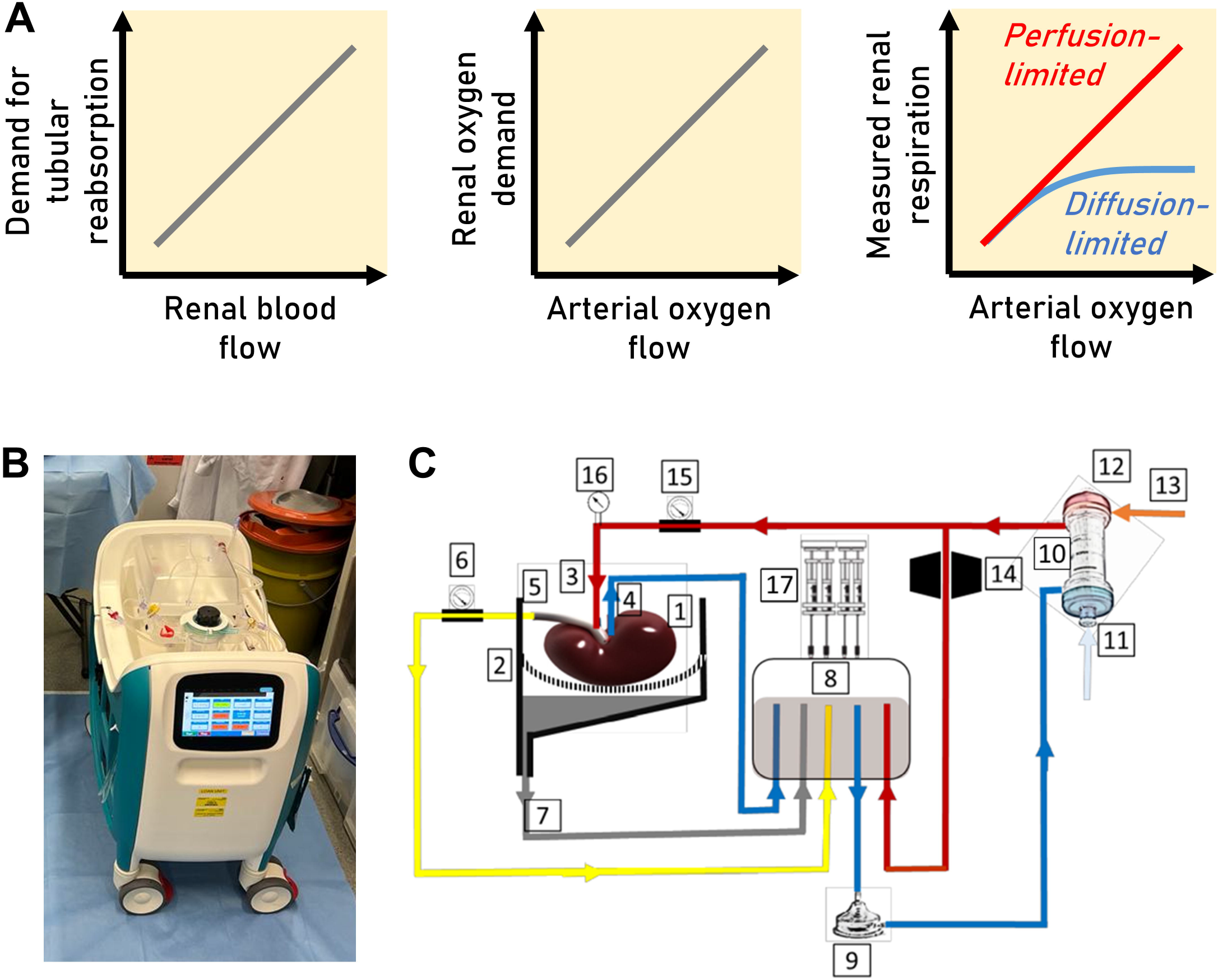
(A) An increase in arterial blood flow increases glomerular filtration and hence increases demand for tubular re-absorption. Thus, unique to the kidney, there is a positive relationship between arterial oxygen flow and renal oxygen demand. If oxygen exchange from blood to the kidney was perfusion-limited, then a linear relationship between arterial oxygen flow and renal respiration is anticipated (red). Otherwise, if oxygen exchange from blood to the kidney was diffusion-limited, then the relationship between arterial oxygen flow and renal respiration is expected to be flatter (blue). Thus, it is possible to test for diffusion-limited oxygen exchange by relating measurements of renal respiration with arterial oxygen flow. (B). Image of machine for normothermic kidney perfusion. (C) Schematic diagram of the circuit in the kidney perfusion machine: 1: kidney, 2: organ containing with perforated kidney sling, 3: arterial cannula at kidney inlet, 4: venous cannula at kidney outlet; 5: ureter outlet duct; 6: urine flow meter; 7: duct for recirculation of fluids leaked by the kidney; 8: soft-shell reservoir; 9: centrifugal perfusion pump; 10: oxygenator and heat exchanger; 11: heat exchanger water inlet; 12: heat exchanger water outlet; 13: oxygenator has inlet; 14: in-line blood gas analysis sensor; 15: arterial flow meter; 16: arterial pressure sensor; 17: infusion or syringe pump.

We find that the renal respiratory rate varies considerably in perfused kidneys and is limited by the kinetic properties of oxygen release from RBCs. Critically, respiratory rate correlated weakly with the classical definition of oxygen delivery based on the product of Q, Hb and S_A_. Consistent with a diffusion-limited process, introducing a term for the rate of oxygen release from RBCs produced a strong positive correlation with renal respiration. Our observations indicate that during capillary transit, oxygen release from RBCs is rate-limiting for oxygen uptake into tissues. The impact of our findings is three-fold. Our results challenge the paradigm that gas exchange is rapid, highlight the need to consider RBC gas-handling kinetics in clinical care, and justify efforts to optimise blood storage regimes.

## RESULTS

### Obtaining kidneys and blood for normothermic perfusion

A total of 30 deceased-donor kidneys, retrieved with the intention of transplantation and allocated to consenting recipients, were transplanted following a period of normothermic machine perfusion (NMP-K). Perfusion was undertaken as part of a phase 1 trial (Normothermic Kidney Perfusion Phase 1, NKP1) designed to evaluate the safety and feasibility of prolonged-duration normothermic machine perfusion of the kidney. Organ retrieval procedures were conducted in accordance with normal clinical practice and kidneys were transported to Oxford under conditions of static cold storage. Following receipt and transfer to the operating room, conventional backtable surgery was performed prior to attachment of the kidney to the investigational device and perfusion at 37°C with RBC-based perfusate prior to transplantation. This clinical trial, and analysis of samples derived from it, was performed in accordance with the principles of the declaration of Helsinki, and received ethical approval from the UK National Health Service (NHS) North West – Greater Manchester South Research Ethics Committee (REC ref. 20/NW/0442) and the Medicines and Healthcare products Regulatory Agency (MHRA). Units of blood (of ABO-Rh type matched to the organ donor) for perfusion were issued by the Blood Bank service at Oxford University Hospitals. All units of blood were subject to conventional collection and storage procedures, with cold-chain maintained up until addition to the perfusion machine. An image of the machine and a schematic diagram are shown in **Figure 1B/C**.

### Stored bloods used for kidney perfusion manifested a range of kinetic dysfunction

Single-cell oxygen saturation imaging was performed to determine the kinetics of oxygen unloading in terms of time constant and capacity. For this experiment, RBCs were loaded with CellTracker DeepRed and Calcein to image red and green fluorescence under superfusion in a microfluidic chamber (**Figure 2A**). Solution exchange in the microfluidic chamber is rapid, and not meaningfully rate-limiting for the biological process of oxygen unloading from RBCs (**Figure 2B**). The ratio of fluorescence describes the time-course of oxygen saturation of haemoglobin, which manifests an oxygen-dependent shift in absorbance spectrum that affects signal on the red channel more than on the green. Oxygen unloading was triggered by exposing RBCs to a 30-second period of anoxia (solution bubbled with 100% N_2_, supplemented with 2 mM dithionite) by means of rapid solution-switching valves supplying in the microfluidic chamber over a microscope objective (**Figure 2C**).

**Figure 2:**
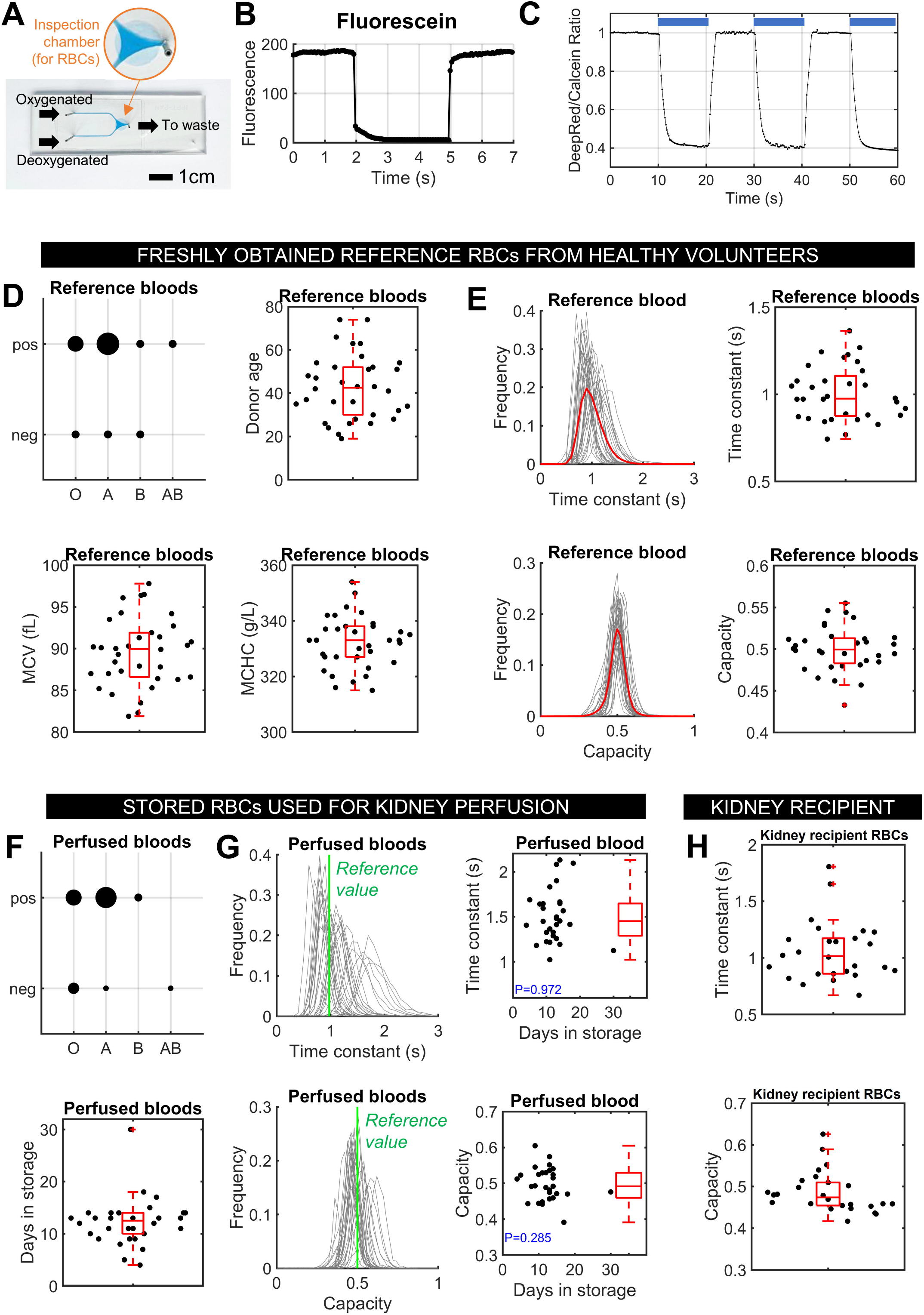
Single-cell oxygen saturation imaging characterises RBC oxygen-handling kinetics. (A) Microfluidic chamber for producing rapid solution exchange in imaged RBCs under superfusion. (B). Rapid exchange is achieved in the millisecond scale (frame acquisition: 51 ms). (C). Experiment on freshly collected venous blood showing time course of oxygen unloading from imaged RBCs, quantified in terms of time constant (*τ*) and capacity (*κ*). (D). Reference bloods freshly obtained from veins of healthy volunteers. Distribution by blood group, donor age, mean corpuscular volume (MCV) and mean corpuscular haemoglobin concentration (MCHC). On each box, the central mark indicates the median, bottom/top edges indicate the 25^th^/75^th^ percentiles, the whiskers extend to the most extreme data points not considered outliers, and outliers are plotted individually. (E). Analysis of reference bloods. Top: time constant frequency distribution (red is mean) and statistics; bottom: capacity frequency distribution (red is mean) and statistics. (F). Stored bloods used for perfusions. Distribution by blood group and distribution of storage time (days). (G). Analysis of stored bloods. Top: time constant frequency distribution (green shows reference mean) and statistics; bottom: capacity frequency distribution (green shows reference mean) and statistics. (H). Analysis of venous blood obtained from recipient of the kidney. Top: time constant statistics; bottom: capacity statistics.

First, a compilation of reference values was obtained for freshly collected venous blood from 32 healthy volunteers (registered blood donors). **Figure 2D** shows the volunteer’s blood group and age (median 42.2 years). Mean corpuscular volume (MCV; median 90 fL) and mean corpuscular haemoglobin concentration (MCHC; median 333 g/L) were within normal levels. O_2_-handling parameters obtained by single-cell oxygen saturation imaging are shown in **Figure 2E**. The median time-constant of unloading was 0.9754 s, consistent with previous recordings^9,23^. The change in fluorescence ratio, described here as capacity, was 0.4994 (where zero would indicate no unloading capacity).

Next, measurements were made on stored bloods used for kidney perfusions. The duration under storage of units varied from 4 to 30 days, with a median of 12.5 days (**Figure 2F**). Stored bloods manifested a range of kinetic behaviours, including those with substantially slower oxygen-unloading compared to reference bloods (**Figure 2G**). The functional impairment incurred during standard storage did not correlate strongly with storage duration. This finding is consistent with a previous observation that the progression of storage lesion is highly donor unit-dependent^9^, which means that storage duration per se is not an accurate proxy of RBC functional quality. Any correlations to organ function must therefore consider a direct measure of oxygen release from RBCs. Venous bloods of the kidney recipient were also analysed for RBC oxygen-unloading and suggested a largely normal range, with two slow outliers (**Figure 2H**).

### Normothermic kidney perfusion generates data on renal function and respiratory rate

The median kidney mass was 227 g (range: 140-410 g) and the duration of normothermic renal perfusion varied from 1h to 23h, with a median of 5h30min. During this time, continuous second-by-second measurements of renal blood flow, temperature, blood pressure, arterial pH, arterial partial pressures of oxygen and carbon dioxide, and urine flow were recorded by the perfusion machine. Regular point-measurements of arterial and venous blood gases, biochemistry, haemoglobin concentration, and saturation were made by means of an external blood gas analyser (Radiometer ABL-90 FLEX). Regular samples of the urine produced during NMP-K were also taken and analysed. **Figure 3A** summarises data on kidney mass, perfusion duration, time-averaged arterial blood flow, arterial [Na^+^], creatinine clearance and arterial Hb. **Figure 3B** presents data on arterial and venous P_O2_, P_CO2_ and calculated O_2_ content. Blood flow over the first 6 hours of perfusion was relatively stable (**Figure 3C**). In contrast, arterial pH tended to start from an acidic value close to 7.1 and gradually recover in the first 3 h of perfusion (**Figure 3D**).

**Figure 3:**
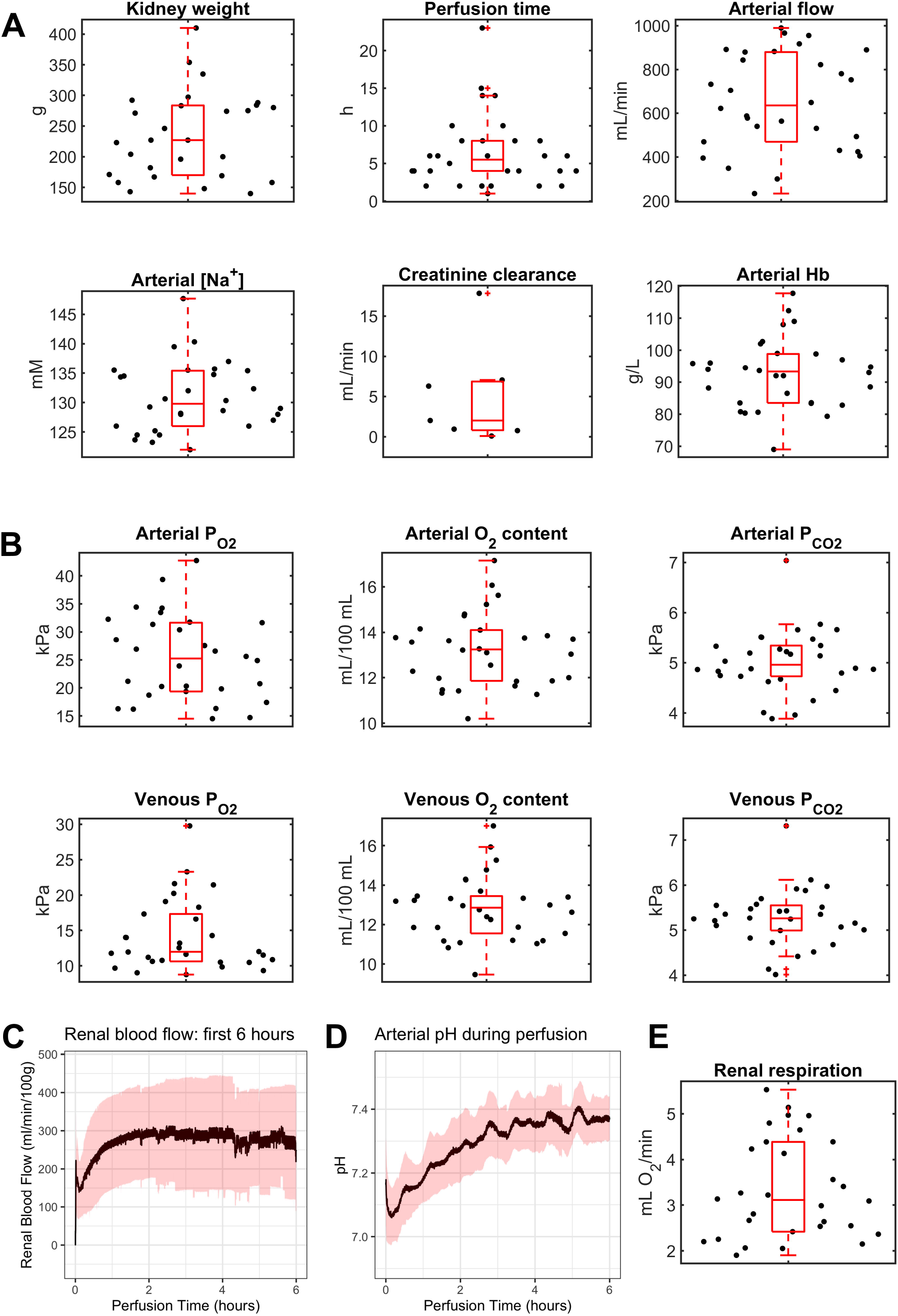
Normothemic kidney perfusion provides comprehensive data on renal function. (A) Data on kidney weight, perfusion time, time-averaged arterial blood flow, arterial [Na^+^], creatinine clearance, and arterial haemoglobin (Hb). (B). Analysis of arterial and venous gases. (C). Time course of blood flow for first 6 h of perfusion. (D). Time course of arterial pH for first 6 h of perfusion. (E). Renal respiration calculated from blood gases.

From these data, it is possible to calculate renal respiratory rate, v′_R,O2_, by mass balance of arterial, venous, and urinary flows of O_2_. Briefly, arterial blood inflow (Q_A_) was assumed to equal the sum of venous blood outflow (Q_V_) and urinary outflow (Q_U_):

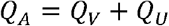

Q_V_ was calculated from measurements of Q_A_ and Q_U_. The oxygen content of arterial blood (mL O_2_/L) was the sum of dissolved oxygen and oxygen held of haemoglobin:

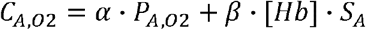

where *α* is O_2_ gas solubility in water, P is partial pressure, *β* is maximal binding capacity for O_2_ per gram of Hb, [Hb] is the concentration of Hb (g/L) after subtracting methaemoglobin and carboxyhaemoglobin, and S_A_ is arterial O_2_ saturation (scaled to 1). Under our experimental conditions, *α* was 0.225 mL O_2_/L blood/kPa and *β* was 1.39 mL O_2_/g Hb. A similar equation holds for venous blood, with a correction to [Hb] that accounts for the concentrating effect of fluid diversion to urine, equal to a factor of Q_A_/Q_V_:

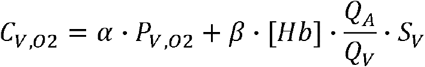

By mass balance, arterial (v′_A,O2_), venous (v′_V,O2_), urinary (v′_U,O2_) and respiratory (v′_R,O2_) O_2_ flows are constrained by the equation:

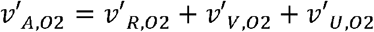

which can be expressed in terms of flows and oxygen content, as derived in the **Appendix**:

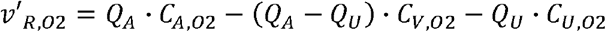

This equation can be re-written in terms of measured parameters and constants:

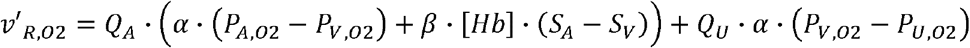

The renal respiratory rate (v′_R,O2_) was calculated at various points during perfusion, ensuring that arterial and venous blood gas measurements were precisely matched. Calculated v′_R,O2_ varied considerably, from 1.9 to 5.5 mL O_2_/min, with median of 3.11 mL O_2_/min (**Figure 3E**). This analysis indicates major organ-to-organ differences in the ability of kidneys to extract oxygen. Differences in v′_R,O2_ were not due to variation in kidney mass (Pearson’s *ρ*=0.10, P=0.24).

### RBCs undergo partial rejuvenation during perfusion but retain a degree of kinetic dysfunction

To relate renal respiration with RBC gas-handling properties, samples of blood were taken during kidney perfusions. Transfusion of stored blood has been shown to have a rejuvenating effect on RBCs *in vivo*^34^. To determine how RBC gas-handling is affected during kidney perfusions, the evolution of unloading time constant (**Figure 4A**) and capacity (**Figure 4B**) were recorded during perfusions. There was an overall trend for RBCs to slow down oxygen release, and then recover. During this time, the pH of blood also showed a time-dependence, gradually recovering from mildly acidotic to physiological levels. Thus, correlation analyses used RBC kinetics once pH had stabilised, labelled as end-point in Figure 4. The distributions of time constant and capacity at end-point are shown on **Figure 4C/D**. Perfusion had a significant accelerating effect on time constant but no significant effect on capacity, as shown in **Figure 4E**.

**Figure 4:**
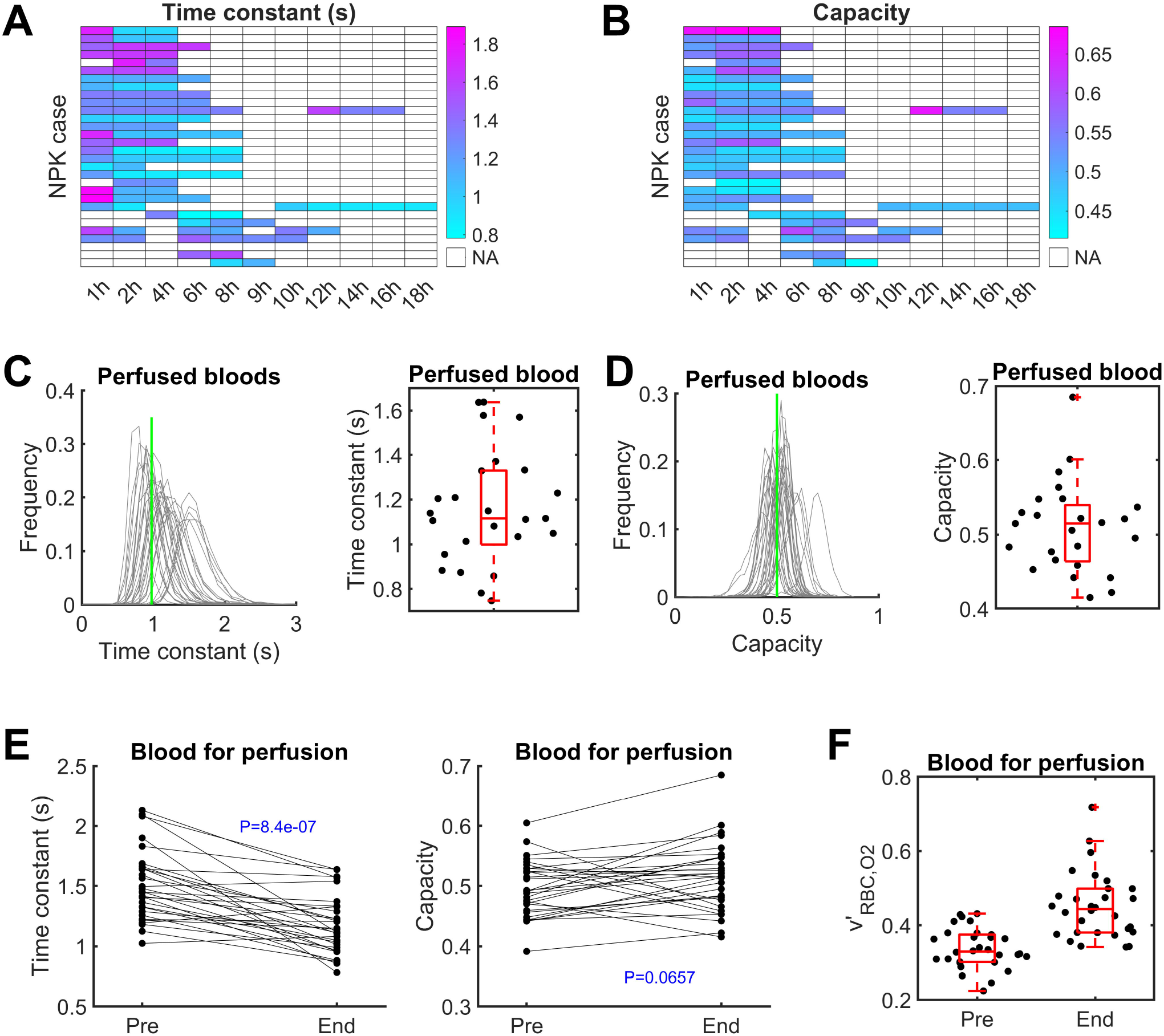
Evolution of changes in RBC oxygen-handling kinetics during kidney perfusion. (A) RBC oxygen unloading time constant at various points (x axis) during perfusion for the 30 cases (y axis). (A) RBC oxygen unloading capacity at various points (x axis) during perfusion for the 30 cases (y axis). (C). Analysis of stored blood at the end of perfusion. Time constant frequency distribution (green shows reference mean). (D). Analysis of stored blood at the end of perfusion. Capacity frequency distribution (green shows reference mean). (E). Effect of kidney perfusion on RBC oxygen unloading time constant (significant decrease; paired t-test) and capacity (no significant change; paired t-test). Pre: packed RBCs prior to perfusion; End: towards end of perfusion. (F). Initial oxygen unloading rate calculated from the time constant and capacity. Pre: packed RBCs prior to perfusion; End: towards end of perfusion.

The ability of RBCs to release oxygen can be quantified in terms of the peak unloading rate, which would take place at the beginning of a capillary. For a diffusion-limited process, this rate would be sufficiently slow to prevent equilibration by the time RBCs reach the venous end of the capillary. Peak unloading rate was estimated from the time course of RBC oxygenation (S_RBC_). The mono-exponential curve that describes the time course of unloading is:

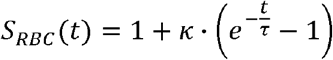

The time-derivative of this relationship is therefore

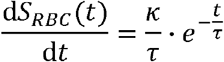

The initial (peak) rate of release from the RBC (v′_RBC,O2_) is given by the ratio of capacity and time constant:

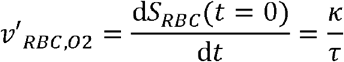

**Figure 4F** plots calculated v′_RBC,O2_ in stored bloods before and during perfusion. The analysis reveals considerable differences in the capacity of RBCs from different blood bags to release oxygen during normothermic perfusion. If gas exchange were diffusion-limited at capillaries, then at least the slower range of these release rates would become rate-limiting for the tissue respiratory rate (v′_R,O2_). This was tested by seeking the most accurate correlation between oxygen delivery and renal respiration.

### Renal respiration correlates with oxygen delivery after factoring oxygen unloading from RBCs

A unique property of the kidney is that blood flow and respiratory rate are coupled under physiological conditions because higher flows produce more tubular filtrate and demand more oxygen for epithelial active transport. Thus, renal respiratory rate (v′_R,O2_) is expected to be directly proportional to arterial oxygen delivery, if the process is perfusion limited, i.e. gas exchange at capillaries is not rate-limiting. However, kidneys in the present study were perfused with RBCs spanning a range of gas-handling ability, including some that fall within the reference range and others substantially slower. If oxygen delivery becomes a diffusion-limited process, particularly for the slower-releasing RBCs, then no proportionality would be observed between arterial oxygen delivery and respiratory rate. To that end, correlation was sought between v′_R,O2_ and the canonical definition of oxygen delivery (D_O2_^PL^) that assumes a perfusion-limited process and is defined as the product of Q_A_ and arterial oxygen content (C_O2,A_)::

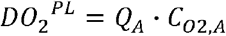

DO_2_^PL^ correlated strongly with arterial blood flow (**Figure 5A**), but it’s relationship with renal respiration (v′_R,O2_) was not statistically significant (Pearson’s *ρ*=+0.36; P>0.05). This finding indicates that oxygen flow carried by arterial blood is a poor predictor of the kidney’s ability to extract oxygen for its respiratory needs. A plausible explanation is that oxygen exchange at capillaries becomes diffusion-limited for at least some stored RBCs, such that the kidney is unable to extract the oxygen that is needed to meet tubular transport demands. In support of this hypothesis, there was a significant negative correlation between oxygen unloading time constant (*τ*) and v′_R,O2_ (**Figure 5C**; Pearson’s *ρ*=-0.535; P=0.00234). This finding suggests an effect of RBC gas-handling on kidney respiration. To test for a diffusion-limited process, correlations were sought between v′_R,O2_ and a modified definition of oxygen delivery that includes a factor describing the initial rate of oxygen unloading from RBCs:

**Figure 5:**
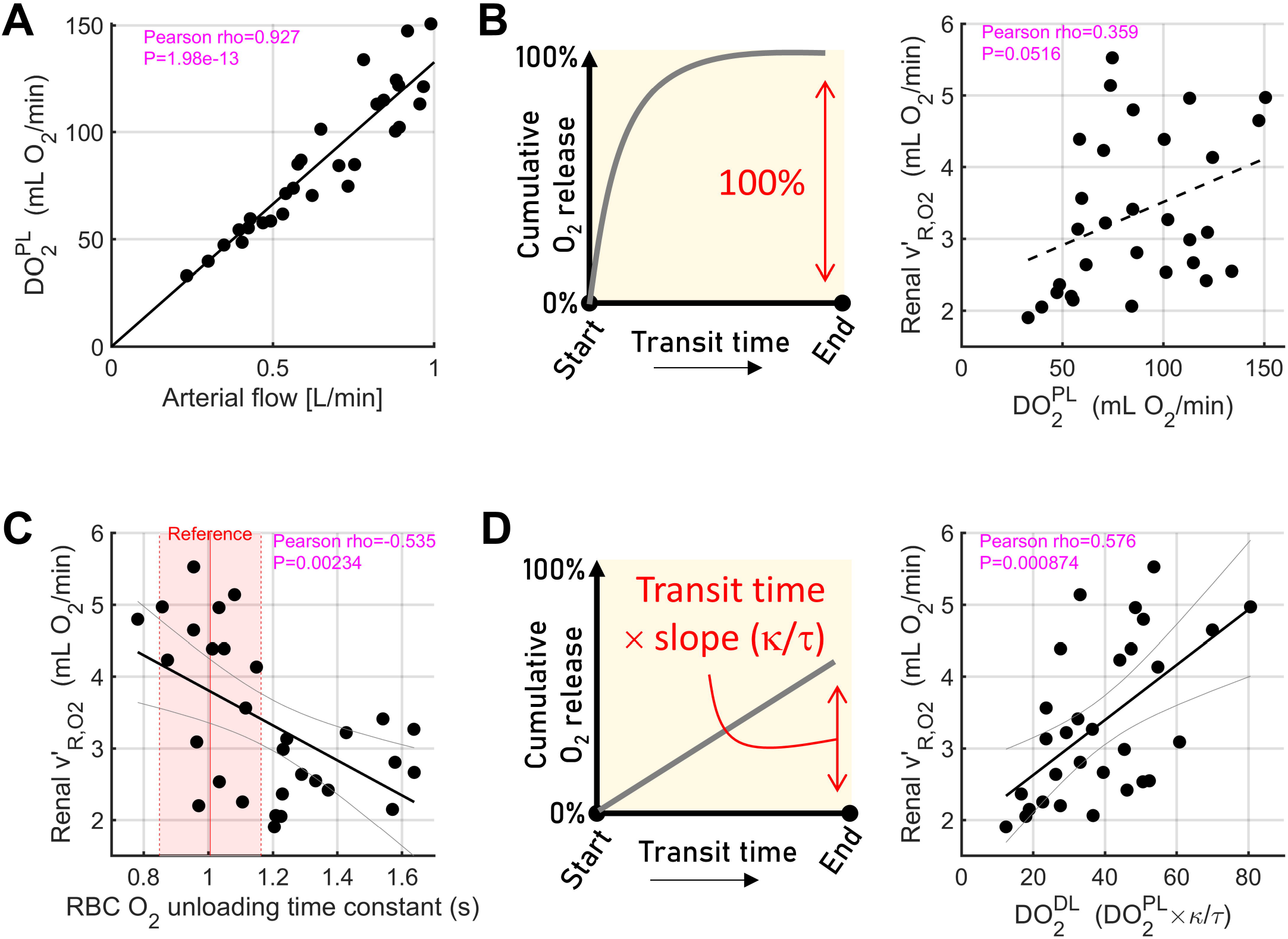
Relating oxygen delivery with renal respiratory rate. (A). Strong linear relationship between arterial blood flow and arterial oxygen delivery calculated assuming perfusion-limited gas exchange. Statistical testing by Pearson’s correlation coefficient. (B). Non-significant correlation between arterial oxygen delivery and renal respiratory rate (v′_R,O2_). (C). Strong, negative correlation between the RBC oxygen unloading time constant and renal respiration. Reference range (red) from reference bloods. (D). Strong, positive correlation between arterial oxygen delivery scaled by RBC initial oxygen unloading rate (*κ*/*τ*) and renal respiration.

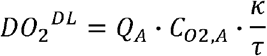

Unlike the canonical definition of oxygen delivery (DO_2_^PL^), the diffusion-limited variant (DO_2_^DL^) produced a strong (Pearson’s *ρ*=+0.58) and statistically significant (P=0.000874) correlation with renal respiration, as shown in **Figure 5D**.

The difference between these two models relates to the ability of respiring tissue to extract oxygen from the blood. Under perfusion-limited delivery, the kidneys will be able to extract the amount of oxygen required by metabolism because of sufficiently rapid exchange between tissues and capillaries. Thus, the initial rate of oxygen release from RBCs becomes irrelevant because full equilibration will occur imminently during capillary transient. Consequently, small changes to this parameter (e.g. associated with the storage lesion) will not affect tissue oxygenation. In a diffusion-limited scenario, however, the kidneys will only extract a fraction of their demand because blood transiting capillaries will not have enough time to reach equilibrium. Consequently, renal respiration will be lower than expected for a given blood flow, which explains the relationship in **Figure 5B**. Our finding, that a strong positive correlation is obtained by introducing a factor describing RBC gas-handling (**Figure 5D**), indicates that gas exchange at capillaries is a diffusion-limited process.

## DISCUSSION

Our observations challenge the canonical view that oxygen exchange at systemic capillaries is perfusion-limited, stemming from the assumption that oxygen release from RBCs is rapid relative to the capillary transit time. Based on our findings, we propose that oxygen delivery to tissues can become diffusion-limited, with far-reaching consequences on clinical care. Previously, we showed that the rate of oxygen unloading from RBCs is slower than prior estimates and could be characterised by a time constant of around one second. Given that capillary transit times are typically no longer than a few seconds^30,31^, our findings raised the possibility of diffusion-limited exchange *in vivo*. Demonstrating such a phenomenon in perfused organs is challenging as it requires a well-controlled system, in which respiratory demand for oxygen can be stimulated and compared to actual oxygen supply. The kidney is an excellent experimental model for studying oxygen exchange because its respiratory demand can be manipulated by varying blood flow, and compared to oxygen consumption measured from arterial and venous gas analyses. At the same time, it is possible to perfuse kidneys with stored blood, spanning a range of kinetic ability, to test if slower-unloading bloods evoke a lower-than-expected respiratory rate. Our results, based on a large number of perfused organs, show unequivocally that renal respiration is not a simple function of arterial oxygen delivery, but requires an additional factor describing oxygen release from RBCs. We conclude that at least for RBCs manifesting a kinetic dysfunction (such as that incurred in storage), oxygen release is a diffusion-limited process.

Our findings have implications on our understanding of gas exchange. Contemporary models of blood transport must now consider the possibility that at least under certain conditions, oxygen release may be diffusion-limited and partial pressures in blood may not have sufficient time to equilibrate with tissue oxygen tension. Oxygen delivery via a diffusion-limited process is less intuitive to predict and may require mathematical models. For example, an increase in blood flow may not improve oxygen release into tissues, which then limits the ability of regulatory processes, such as cardiac output and vascular flow redistribution, to meet respiratory demands. A diffusion-limited process may result in an oxygen debt, and the only way of improving delivery would be to address the rate-limiting process underpinning slow gas exchange in capillaries, such as dysfunctional RBCs.

Clinical care guidelines will need to consider the impact of RBC function when seeking ways to improve tissue oxygenation. In case of severely diffusion-limited gas exchange, tissue oxygenation will not respond to inotropes, as this will merely abbreviate transit time and reduce fractional release. Attempts to raise Hb levels will need to consider the kinetic quality of the transfused product. For example, a transfusion of kinetically dysfunctional RBCs in units that underwent severe storage lesion, may not produce favourable outcomes as this introduces cells that are unable to release oxygen fully, despite an increase in oxygen-carrying capacity. Diseases that reduce oxygen release from RBCs, such as hereditary spherocytosis (HS), may result in more severe diffusion-limited oxygen delivery to tissues. One intervention for HS is partial or full splenectomy, as this has been shown to raise haematocrit by reducing the rate at which spherocytes are removed from circulation. However, splenectomy achieves a higher haematocrit by enriching blood with kinetically-slower cells, and the overall outcome may not necessarily be beneficial to the patient.

Our findings highlight the importance of monitoring stored bloods for their kinetic quality. We have previously shown that the rate of kinetic attrition varies considerably between donor units, and may relate to the donor or way in which bloods are processed^9^. Storage duration is a poor proxy of kinetic quality, and therefore should be avoided when deciding how to match blood bag with recipient or when randomising blood bags for clinical trials studying transfusion efficacy. Instead, we propose that RBC quality should be monitored, and when necessary, rejuvenated biochemically to ensure that recipient outcomes are not compromised by problems associated with diffusion-limited gas exchange.

A limitation of our study is that it was limited to donated kidneys. It would be important to study the impact of rate-limiting gas exchange on other organs such as the heart, brain, and skeletal muscle, where it is imperative that oxygen delivery is as high as possible. Nonetheless, the kidney is an excellent model for studying gas exchange because of its unique physiological properties, but also its essential role in the body. Future trials should investigate how different storage protocols or rejuvenation techniques improve outcomes, and for methods that can distinguish kinetically dysfunctional blood units so that these can be matched to the correct recipient. For instance, rapid kinetics will be more critical for patients in the setting of major haemorrhage, or in clinical ex-vivo organ perfusion ahead of transplant, where the imperative is to restore oxygenation immediately. In contrast, the efficacy of top-up transfusions in anaemic patients is less likely to be influenced by gas-handling ability. One parameter that may inform RBC kinetic quality is flow cytometric side-scatter^9^.

In summary, we provide evidence in perfused human kidneys that oxygen delivery can be diffusion-limited when RBCs undergo a modest kinetic defect associated with storage. Diffusion-limited gas exchange has implications on clinical care and blood banking.

## ONLINE METHODS

### Approvals

The UK National Health Service (NHS) North West – Greater Manchester South Research Ethics Committee gave ethical approval for the work (REC ref. 20/NW/0442).

### Reference bloods

Bloods for obtaining reference data on oxygen-handling were obtained from NHS Blood and Transplant (registered customer number T361). Healthy volunteers were donors registered with the blood service, who consent for small samples of blood to be studied for research purposes. No bloods were stored beyond immediate measurements and all samples were destroyed after measurements.

### Single-cell oxygen saturation imaging

The kinetics of oxygen unloading from RBCs were studied using a modification of a previously published fluorescence imaging method^23^ adapted for a microfluidic chamber^29^. An revised design of microfluidic chamber was used, as described below. Briefly, an aliquot of blood was diluted in Hepes-buffered normal Tyrode (NT; pH 7.4 at RT), and RBCs were loaded with CellTracker DeepRed and Calcein. After allowing 10 min for dye uptake, cells were applied to the microfluidic chamber pre-coated with poly-L-lysine and allowed to settle for another 5 mins. Once attached to the coverslip surface, valves were alternated every 10 s to expose cells to a normoxic stream of NT (equilibrated with air) or an anoxic stream (bubbled with N_2_, supplemented with 2mM dithionite). The time course of oxygen unloading was fitted with a monoexponential curve that describe the time constant and overall change. The time course of oxygen unloading in individual cells were fitted with a mono-exponential curves that described the time constant and overall change. Experiments were repeated at least 3 times (independent loadings). At least 10 fields of view were obtained per sample.

### Microfluidic chamber for rapid solution exchanges

The chamber consisted of two input lines and one output. The input channels are placed symmetrically to the central axis of the device. The input ducts of an initial width of about 400μm approach the main chamber of the device at an angle of 45 degrees to the axis, gradually increasing width and smoothly transiting to the common central chamber. Such a regular and even design prevents flow irregularity. The output of the chamber is placed on the axis of the device to ensure symmetry. The microfluidic structure was fabricated in a transparent polycarbonate plate (Makrolon® GP, Bayer, Germany) by milling and then closed by attaching a cover slip forming a glass bottom surface for maintaining RBC in the observation chamber. Milling was performed using a CNC machine (MFG4025P, Ergwind, Gdansk, Poland). A 2-flute fishtail milling component of diameter of 385 μm (FR208, InGraph, Goleniow, Poland) were used for the engraving of the structure of the device. A high-pressure water washer removed turnings and loosely bound bulk material formed during the milling process (Karcher, K7 Premium, Winnenden, Germany). Chambers were then washed with isopropanol and deionized water and dried with compressed air.

## Data Availability

All data produced in the present study are available upon reasonable request to the authors

## APPENDIX

Mass balance of oxygen flow:

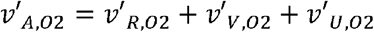

Expanding oxygen flows in terms of fluid flow and its oxygen content:

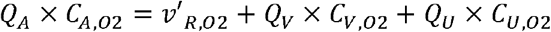

Expressing Q_V_ in terms of Q_A_ and Q_U_:

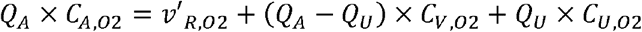

Expanding definitions of oxygen content:

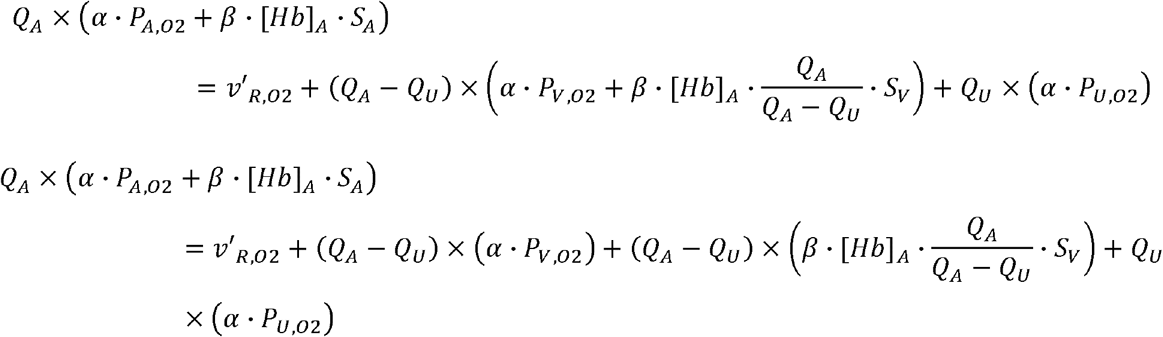

Simplify:

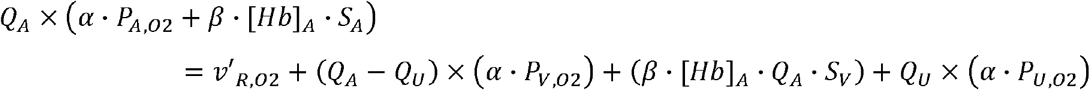

Re-arranging in terms of respiratory rate:

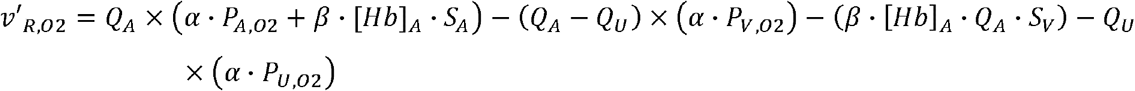

Expand and collect under factors of Q_A_ and Q_U_:

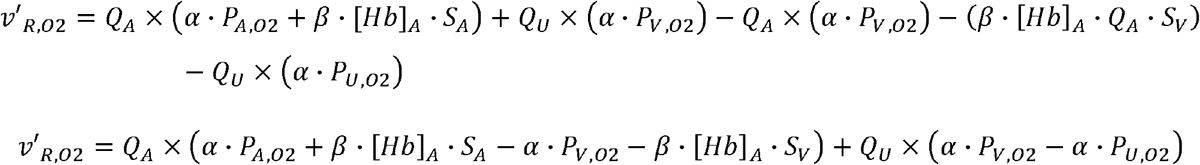

Simplify and collect factors of *α* and *β*:

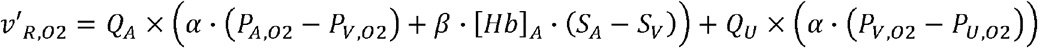

## Notes

### Competing Interest Statement

The authors have declared no competing interest.

### Funding Statement

This study was funded by NIHR and UKRI

### Author Declarations

The UK National Health Service (NHS) North West Greater Manchester South Research Ethics Committee gave ethical approval for the work (REC ref. 20/NW/0442).

